# Projected epidemiological consequences of the Omicron SARS-CoV-2 variant in England, December 2021 to April 2022

**DOI:** 10.1101/2021.12.15.21267858

**Authors:** Rosanna C. Barnard, Nicholas G. Davies, Carl A. B. Pearson, Mark Jit, W. John Edmunds

**Affiliations:** Centre for Mathematical Modelling of Infectious Diseases, London School of Hygiene & Tropical Medicine, Keppel Street, London, WC1E 7HT, UK; South African DSI-NRF Centre of Excellence in Epidemiological Modelling and Analysis, Stellenbosch University, Stellenbosch, Republic of South Africa

## Abstract

The Omicron B.1.1.529 SARS-CoV-2 variant was first detected in late November 2021 and has since spread to multiple countries worldwide. We model the potential consequences of the Omicron variant on SARS-CoV-2 transmission and health outcomes in England between December 2021 and April 2022, using a deterministic compartmental model fitted to epidemiological data from March 2020 onwards. Because of uncertainty around the characteristics of Omicron, we explore scenarios varying the extent of Omicron’s immune escape and the effectiveness of COVID-19 booster vaccinations against Omicron, assuming the level of Omicron’s transmissibility relative to Delta to match the growth in observed S gene target failure data in England. We consider strategies for the re-introduction of control measures in response to projected surges in transmission, as well as scenarios varying the uptake and speed of COVID-19 booster vaccinations and the rate of Omicron’s introduction into the population. These results suggest that Omicron has the potential to cause substantial surges in cases, hospital admissions and deaths in populations with high levels of immunity, including England. The reintroduction of additional non-pharmaceutical interventions may be required to prevent hospital admissions exceeding the levels seen in England during the previous peak in winter 2020–2021.

## Introduction

The B.1.1.529 SARS-CoV-2 lineage was first reported to the World Health Organization by South Africa on 24th November 2021, and was designated as the Omicron variant of concern two days later (8). Recent increases in COVID-19 cases and hospitalisations in Gauteng province, South Africa (**Fig. 1a**), along with evidence suggesting Omicron possesses an increased risk of reinfection (9), suggests that populations with high levels of immunity derived from prior infection and/or vaccination may experience new waves of transmission. The Omicron variant possesses a number of concerning mutations (8), with the European Centre for Disease Prevention and Control estimating that Omicron has the potential to cause more than half of all SARS-CoV-2 infections in Europe within the next few months (10). Just over two weeks after its initial detection, Omicron sequences have been reported in 41 countries worldwide (11). There is therefore an urgent need to understand the potential impact of Omicron on COVID-19 disease burden and the mitigating effect of control measures in highly immune populations.

**Figure 1.**
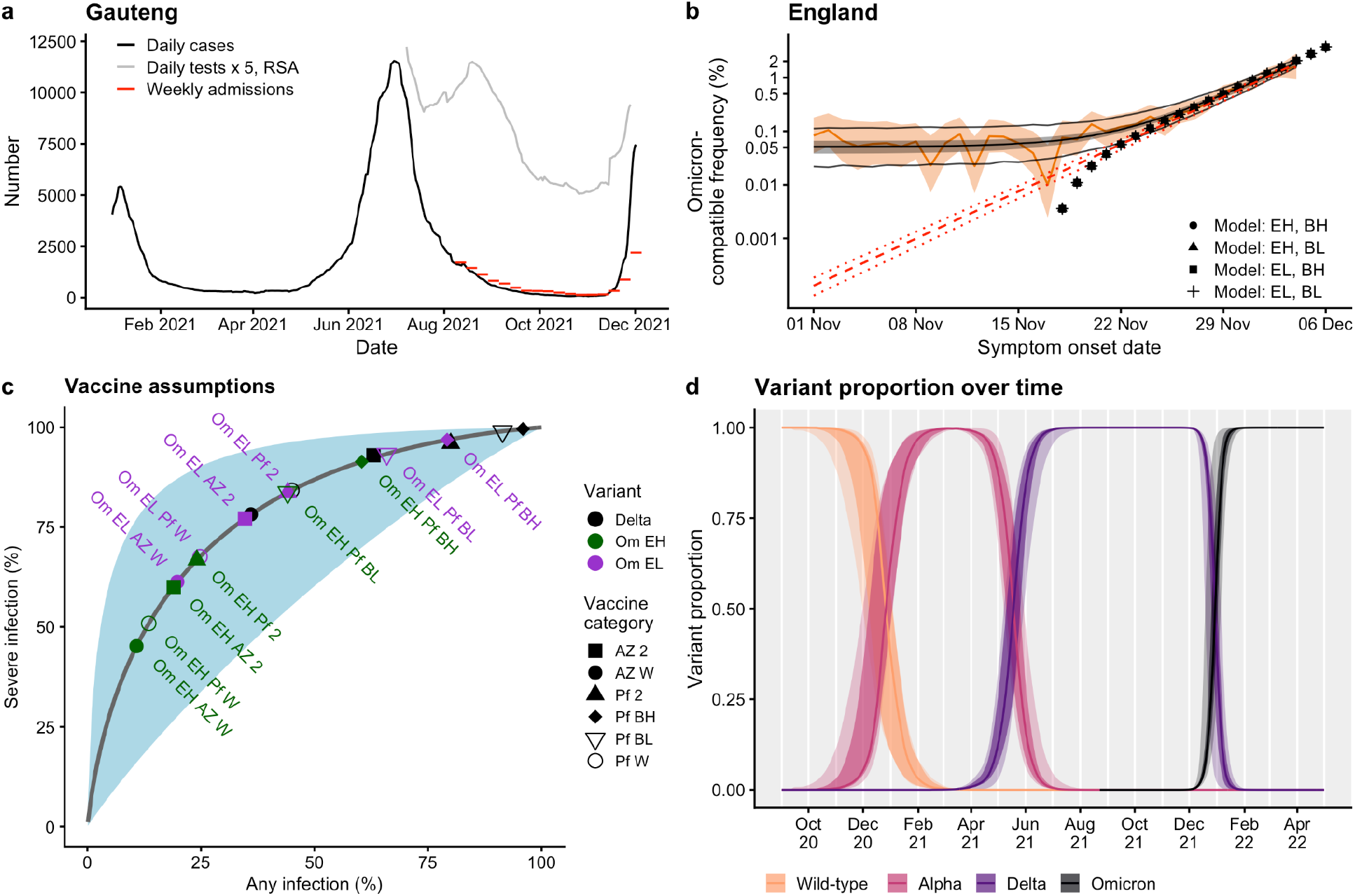
Omicron epidemiology and assumptions. **(a)** Daily cases (black) and weekly hospital admissions (red) for Gauteng province in the Republic of South Africa (RSA), and 1/5 times the number of daily tests recorded in RSA (grey). **(b)** Proportion of symptomatic Pillar 2 cases with S gene target failure (SGTF), a proxy for the Omicron variant in England, point estimate (orange line) and 95% binomial confidence intervals (orange ribbon); statistical fit to proportion of symptomatic Pillar 2 cases with SGTF (central black line, median; grey ribbon, 95% credible interval; outer black lines, 95% credible interval of expected sampling variability); statistical fit to estimated true proportion of Omicron variant over time assuming constant logistic growth, median and 95% confidence intervals (red dashed lines); transmission model proportion of Omicron shown for four primary scenarios (black points). EH: high immune escape; EL: low immune escape; BH: high booster efficacy; BL: low booster efficacy. Logistic scale on y-axis. **(c)** The modelled relationship between vaccine effectiveness against any infection (x-axis) versus severe infection (y-axis), from Khoury et al. Figure 3a (7). Assumed values for vaccine effectiveness against infection (x-axis) and hospitalisation (y-axis) for the AstraZeneca (AZ) and Pfizer and Moderna (Pf) vaccines are also plotted (2: dose 2, W: waned, BH: boosters high, BL: boosters low), for the Delta (black) and Omicron variants. Om EL (purple) corresponds to the low immune escape Omicron scenario (a 5.1-fold drop in neutralisation titre between Delta and Omicron), whilst Om EH (green) corresponds to the high immune escape Omicron scenario (a 12.8-fold drop). **(d)** The modelled proportion of SARS-CoV-2 variants in England over time, from September 2020 until April 2022: from left to right, wild-type, Alpha B.1.1.7, Delta B.1.617.2 and Omicron B.1.1.529 SARS-CoV-2 variants.

One such example is England, which achieved 81% two-dose vaccine coverage in the eligible (aged 12 and over) population by 1st December 2021 (12), and has also experienced three large COVID-19 waves. In England, the first Omicron cases were confirmed on 27th November 2021 (13), with more than 100 cases reported by 3rd December 2021 (14) and 1139 cases as of 10th December 2021 (15). The prevalence of S gene target failure (SGTF), a proxy for the Omicron variant (8), has risen sharply (**Fig. 1b**), suggesting that more Omicron cases are likely to be reported. In response to concern about Omicron, the UK government announced limited additional precautionary control measures on 27th November 2021 (16) and widened and accelerated the COVID-19 booster vaccination rollout (17), before announcing more extensive “Plan B” measures on 8th December 2021 (18). We adapt an existing mathematical model of SARS-CoV-2 transmission (19) to generate scenarios outlining the potential impact of Omicron in England and the effect of different control measures.

## Methods

Neutralisation studies on Omicron are ongoing, so we rely on a combination of estimates of the fold reduction in neutralisation titre for previous variants of concern (VOCs) as well as early studies explicitly considering Omicron neutralisation (1–5) to inform our assumptions about the level of immune escape Omicron might possess. The largest drop in neutralisation titre (8.8 fold) was estimated for the Beta VOC (20), with a 3.9 fold reduction for the Delta variant, compared to the ancestral SARS-CoV-2 virus. We consider two scenarios for the immune escape of Omicron relative to Delta: 5.1-fold (escape low, EL) and 12.8-fold (escape high, EH) reductions compared to our existing assumptions for Delta (19). Since the Delta variant was estimated to have a 3.9 fold reduction in neutralisation compared to the ancestral SARS-CoV-2 virus, these assumptions correspond to overall fold reductions of approximately 20 and 50-fold between the ancestral SARS-CoV-2 virus and the Omicron variant.

We use the relationship between mean neutralisation titre and protective efficacy from Khoury et al. (7) to arrive at assumptions for vaccine efficacy against infection with Omicron, given each drop in neutralisation. We then use Khoury et al.’s modelled relationship between efficacy against any infection and efficacy against severe infection to generate vaccine effectiveness estimates against severe outcomes (**Fig. 1c**). For the effectiveness of booster vaccinations against Omicron, we base two main scenarios on two studies which measured increases in neutralisation titres following the second dose of the primary vaccination course and after booster vaccinations with the Moderna (2.5-fold) and Sinovac (4.9-fold) vaccines (20). We assume that protection against infection for individuals who have received a primary course of the AstraZeneca COVID-19 vaccine before being boosted with either full-dose Pfizer or half-dose Moderna (the current policy in England) is initially increased to the same levels as Pfizer/Moderna, before using the relationship in **Fig. 1c** to scale protection against infection to protection against severe outcomes (hospitalisation and death). We assume that individuals in the recovered disease state who have previously been infected with SARS-CoV-2 have the same level of protection against Omicron as individuals who have received two doses of Pfizer/Moderna. Our estimates for vaccine protection against different outcomes for the Delta variant and for the various Omicron scenarios are shown in **Table 1**. These estimates are broadly in line with early vaccine effectiveness estimates against Omicron and Delta from the UK’s Health Security Agency (UKHSA) (21), with the exception that our assumptions for dose 2 AstraZeneca vaccine protection against Omicron disease are high in comparison (we assume 38.2% and 23.3% protection against disease, whereas the UKHSA study’s highest estimate for protection against Omicron disease with two doses of AstraZeneca is 5.9%).

**Table 1.** Vaccine efficacy assumptions. Overall vaccine efficacy (%) against infection, symptomatic disease, hospitalisation, and mortality, and conditional vaccine efficacy (%) against transmission given breakthrough infection, for the Delta and Omicron variants (low escape scenario versus high immune escape scenarios listed separately) and for AstraZeneca versus Pfizer/Moderna primary course in the model. In England, individuals who receive COVID-19 booster vaccinations receive a full dose of Pfizer or a half dose of Moderna, regardless of which primary course was received. AZ = ChAdOx1 vaccine; mRNA = BNT162b2 or mRNA-1273; 1 = one dose; 2 = two doses; BL = booster dose, low booster efficacy scenario; BH = booster dose, high booster efficacy scenario; W = waned from vaccine protection.

| Variant | Outcome | AZ primary, mRNA booster |  |  |  |  | mRNA primary, mRNA booster |  |  |  |  |
| --- | --- | --- | --- | --- | --- | --- | --- | --- | --- | --- | --- |
|  |  | 1 | 2 | BL | BH | W | 1 | 2 | BL | BH | W |
| Delta | Infection | 43.0 | 63.0 | 91.4 | 95.9 | 36.0 | 62.0 | 80.0 | 91.4 | 95.9 | 45.0 |
|  | Symptomatic disease | 52.0 | 65.0 | 91.9 | 96.1 | 49.0 | 62.0 | 81.0 | 91.9 | 96.1 | 61.0 |
|  | Hospitalisation | 84.0 | 93.0 | 99.0 | 99.6 | 78.2 | 92.0 | 96.0 | 99.0 | 99.6 | 84.2 |
|  | Mortality | 95.0 | 95.0 | 99.0 | 99.6 | 78.2 | 92.0 | 96.0 | 99.0 | 99.6 | 84.2 |
|  | Transmission | 5.0 | 27.0 | 37.0 | 37.0 | 16.5 | 24.0 | 37.0 | 37.0 | 37.0 | 24.0 |
| Omicron (Low escape) | Infection | 23.7 | 34.7 | 65.9 | 79.2 | 19.8 | 34.2 | 44.1 | 65.9 | 79.2 | 24.8 |
|  | Symptomatic disease | 35.7 | 38.2 | 67.6 | 80.3 | 36.1 | 34.2 | 46.9 | 67.6 | 80.3 | 46.7 |
|  | Hospitalisation | 66.3 | 77.1 | 93.3 | 96.9 | 61.3 | 76.7 | 83.7 | 93.3 | 96.9 | 67.6 |
|  | Mortality | 66.3 | 77.1 | 93.3 | 96.9 | 61.3 | 76.7 | 83.7 | 93.3 | 96.9 | 67.6 |
|  | Transmission | 5.0 | 27.0 | 37.0 | 37.0 | 16.5 | 24.0 | 37.0 | 37.0 | 37.0 | 24.0 |
| Omicron (High escape) | Infection | 12.9 | 19.0 | 44.1 | 60.4 | 10.8 | 18.7 | 24.1 | 44.1 | 60.4 | 13.5 |
|  | Symptomatic disease | 26.7 | 23.3 | 46.9 | 62.3 | 28.9 | 18.7 | 27.9 | 46.9 | 62.3 | 38.7 |
|  | Hospitalisation | 49.7 | 60.0 | 83.7 | 91.4 | 45.2 | 59.6 | 66.8 | 83.7 | 91.4 | 50.9 |
|  | Mortality | 49.7 | 60.0 | 83.7 | 91.4 | 45.2 | 59.6 | 66.8 | 83.7 | 91.4 | 50.9 |
|  | Transmission | 5.0 | 27.0 | 37.0 | 37.0 | 16.5 | 24.0 | 37.0 | 37.0 | 37.0 | 24.0 |

We use an existing three-variant model describing transmission of SARS-CoV-2 in England fitted to historic data up to 1st December 2021 (**Fig. 2a**; **Fig. S2**, Supplementary Material) on COVID-19 deaths, hospitalisations, hospital bed and intensive care unit (ICU) bed occupancy, PCR prevalence and seroprevalence, as well as SGTF data describing the introduction of Alpha B.1.1.7, genomic sequencing data describing the introduction of Delta B.1.617.2, and COVID-19 vaccine coverage (19). Initially, the first model variant considers wild-type and other SARS-CoV-2 variants which were circulating in England in 2020. The second model variant considers the Alpha B.1.1.7 VOC from late 2020 and the third variant considers the Delta B.1.617.2 VOC in 2021. Prior to Omicron’s modelled introduction to England, the first variant in the model is switched to the Omicron variant and all parameters are updated accordingly. Any individuals who were recovered from the original first variant are moved into the recovered state from the second variant. Given four combinations of immune escape and booster effectiveness outlined above, we select introduction times, rates of introduction and transmissibility of the Omicron variant relative to Delta in order to match model projections to the observed growth in SGTF cases in England (**Fig. 1b**). Over the whole time series of the epidemic and projecting forwards, this allows us to model the changing proportions of the wildtype, Alpha, Delta and Omicron variants in England (**Fig. 1d**). These results suggest that Omicron will outcompete the Delta variant in England within a matter of weeks (**Fig. 1d**). We assume that Omicron and Delta have identical baseline severity (i.e., severity of infection in an individual with no vaccine or natural infection), but we note that because Omicron is an escape variant, it infects more individuals with pre-existing immunity than Delta and accordingly the average severity per infection is lower than the baseline severity. We also assume that future levels of mobility remain unchanged from the most recent data up to 30th November 2021. All scenarios considered assume a 7.5% reduction in transmission following the introduction of limited mask-wearing measures by the UK government on 30th November 2021, which we assume lasts until 30th April 2022. This is in keeping with our previous estimates for the impact of increased mask-wearing on transmission (22). We further assume that working from home guidance and certification for entertainment venues take effect from 12th December 2021, in line with the announcement of these measures by the UK government on 8th December 2021 (18).

**Figure 2.**
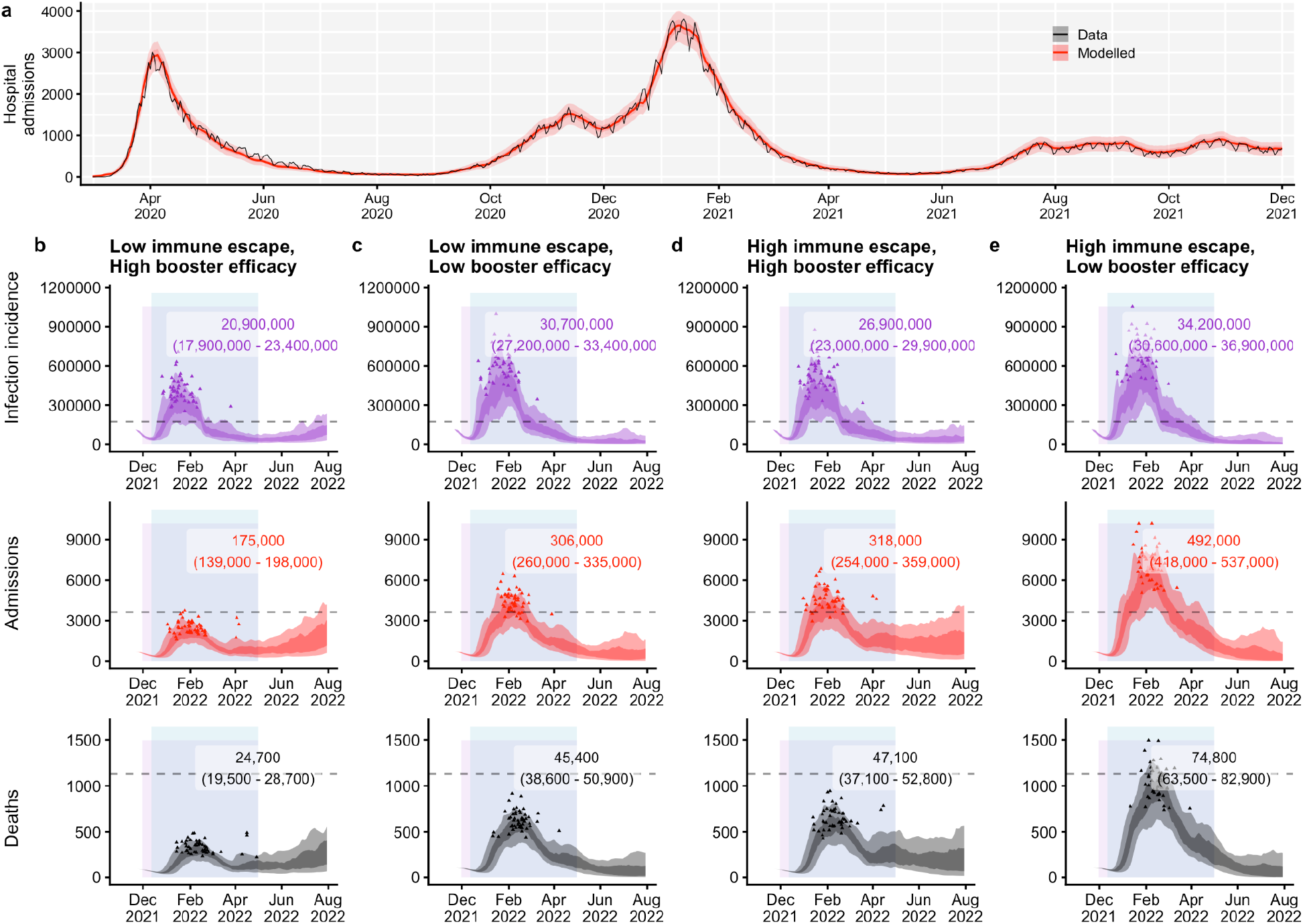
Epidemic scenarios under no further control measures. **(a)** Transmission model fit to hospital admissions data in England, March 2020 – November 2021. **(b–e)** Daily infections, hospital admissions, and deaths projected for an Omicron epidemic in England, with increased mask-wearing from 30th November 2021 to 30th April 2022 (lavender shaded rectangle) and “Plan B” restrictions from 12th December 2021 to 30th April 2022 (blue shaded rectangle). In each panel, the dashed horizontal line shows the previous peak reached during the January 2021 wave in England; the shaded areas show 50% and 90% quantiles across each day, while the small triangles demarcate peaks from each of 50 individual model runs reached between 1st December 2021 and 30th April 2022. Numbers in overlays show the median and 95% projection intervals for total infections, hospital admissions, and deaths between 1st December 2021 and 30th April 2022. **(b)** Low immune escape for Omicron, high booster efficacy. **(c)** Low immune escape for Omicron, low booster efficacy. **(d)** High immune escape for Omicron, high booster efficacy. **(e)** High immune escape for Omicron, low booster efficacy.

## Results

The growth rate of the Omicron variant is determined by two factors: its level of immune escape and its intrinsic transmissibility. The immune escape of Omicron is fixed in our model according to our assumptions of a 5.1-fold to 12.8-fold reduction in neutralisation relative to Delta, as described in the Methods. Given these levels of immune escape, we calibrate the spread of Omicron in our model to data on the growing proportion of SGTF over time in England (**Fig. 1b**), using methods developed previously for analysing the spread of the Alpha variant (23). We estimate that, for a 12.8-fold reduction in neutralisation relative to Delta, Omicron exhibits a 5–10% lower transmission rate than Delta, while for a 5.1-fold reduction in neutralisation relative to Delta, Omicron exhibits a 30–35% higher transmission rate than Delta. These estimates are comparable to estimates made by Pearson et al. for immune escape and transmissibility of Omicron in South Africa (24). We estimate that the Omicron variant is growing in England at an exponential growth rate of *r* = 0.29 per day. This corresponds to a 2.4-day doubling time and a reproduction number *R*_*t*_ = 4.0, assuming a generation interval of 5.5 days with standard deviation 1.8 days (25,26). If the generation interval of Omicron is shorter than 5.5 days, then *R*_*t*_ would be correspondingly lower.

Given the estimated relative transmissibility of Omicron, four immune escape and booster effectiveness scenarios, we begin by projecting SARS-CoV-2 transmission in England from 1st December 2021 until 31st July 2022, under an assumption that no further control measures are introduced except for the policy of mask-wearing in shops and on public transport which started on 30th November 2021, and the introduction of “Plan B” mitigation measures from 12th December 2021 (**Fig. 2**). We model the impact of mask-wearing as a 7.5% reduction in transmission, following our earlier estimate of its potential impact (22). “Plan B” measures comprise: certification requirements for entry to nightclubs or indoor venues with large crowds, modelled as a 17% increase in vaccination uptake for individuals aged 18–29 (22); work from home guidance, modelled by returning work mobility indices to their measured value from the week of 15th March 2021 (following Step 1 of the roadmap out of lockdown (27)); and a face covering mandate, which we assume has no additional effect over and above the separate face covering mandate in effect from 30th November 2021.

All four scenarios project a surge in SARS-CoV-2 transmission beginning in late December 2021, with infections exceeding peak levels recorded during the January 2021 wave in England. Under an assumption of low immune escape, the high booster effectiveness scenario is projected to lead to 175,000 (95% CI: 139,000–198,000) hospitalisations and 24,700 (19,500–28,700) deaths between 1st December 2021 and 30th April 2022 (**Fig. 2b**); the low booster effectiveness scenario projects 306,000 (260,000–335,000) hospitalisations and 45,400 (38,600–50,900) deaths over the same time period (**Fig. 2c**). Under an assumption of high immune escape, the high booster effectiveness scenario projects 318,000 (254,000–359,000) hospitalisations and 47,100 (37,100–52,800) deaths over this period (**Fig. 2d**), whereas the low booster effectiveness scenario projects 492,000 (418,000–537,000) hospitalisations and 74,800 (63,500–82,900) deaths (**Fig. 2e**).

The most optimistic scenario (low immune escape, high booster efficacy, **Fig. 2b**) is projected to lead to peak daily hospital admissions around 60% as high as the peak in January 2021 in England (median peak of 2410 daily admissions, 95% projection interval peak of 1760–3570 daily admissions, compared to 3,800 in January 2021). The most pessimistic scenario is projected to peak at approximately twice the height (7190 [5230–10,040] daily admissions) of the January 2021 peak. The other two scenarios project peaks in hospital admissions between these two extremes, at 4350 (3160–6190) for the low immune escape, low booster efficacy scenario and 4500 (3280–6540) for the high immune escape, high booster efficacy scenario.

We carry forward the most optimistic (low immune escape, high booster efficacy) and most pessimistic (high immune escape, low booster efficacy) scenarios to explore the effect of implementing control measures in addition to the face covering policy in effect from 30th November 2021 (**Fig. 3**). Each roadmap step control measure scenario returns mobility rates to the levels recorded in Google Community mobility data during the equivalent control measure in early 2021, although when modelling roadmap step 2, we assume workplace mobility will remain at the lower level associated with “Plan B” (**Fig. 3a**).

**Figure 3.**
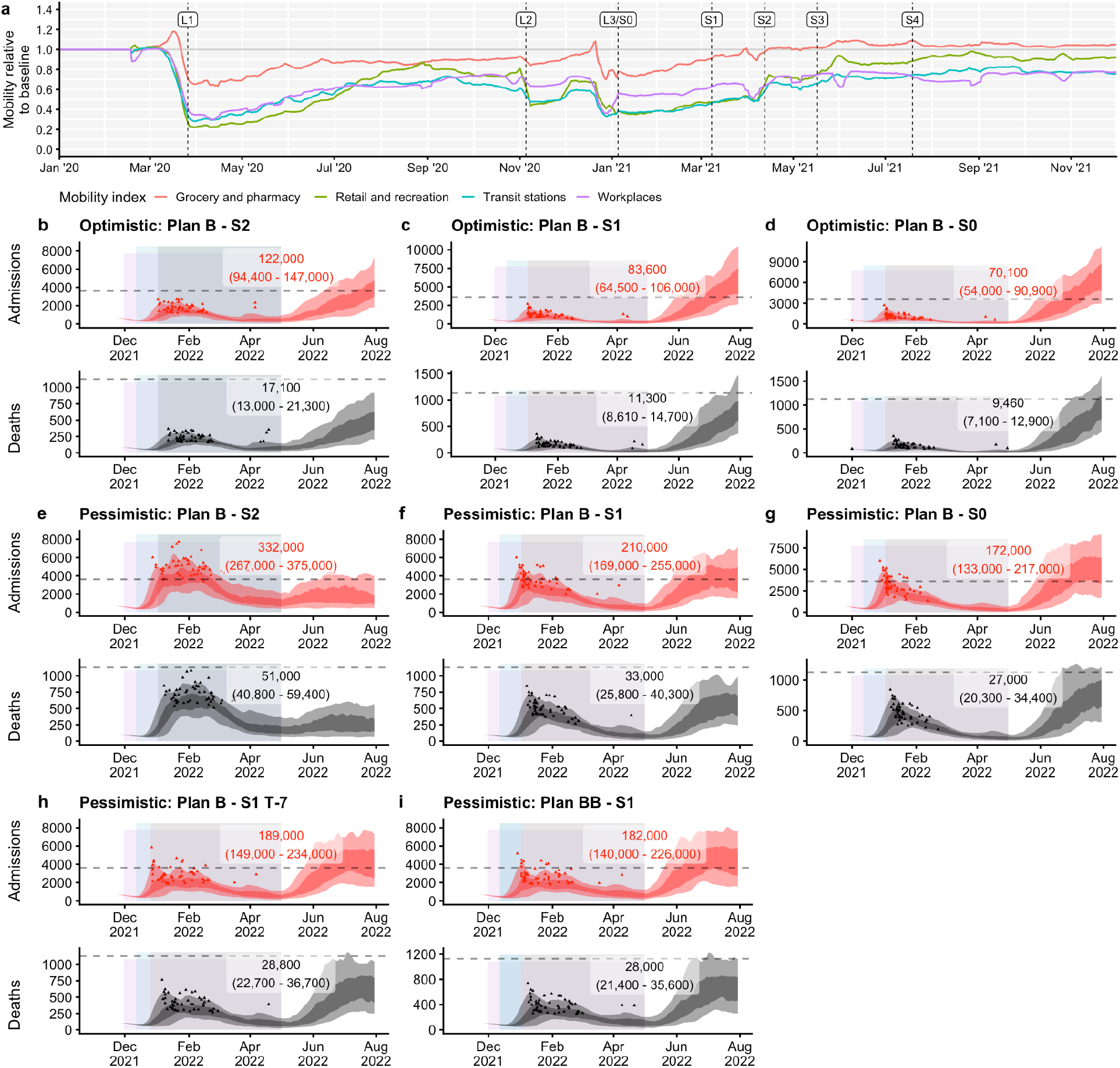
Control measures. **(a)** Google Community mobility data from January 2020 to November 2021, with grocery and pharmacy, retail and recreation, transit stations and workplace mobility indices shown relative to their baseline levels recorded prior to the COVID-19 pandemic. The beginning of each national lockdown and roadmap step is marked with a vertical dashed line (L, lockdown; S, roadmap step). **(b-i)** Daily hospital admissions and deaths projected for a mitigated Omicron epidemic in England, with increased mask-wearing from 30th November 2021 to 30th April 2022 (lavender shaded rectangles), “Plan B” measures (blue shaded rectangles) from 15th December 2021, and a return to various stages of the 2021 roadmap out of lockdown (grey shaded rectangles), implemented from 2nd January 2022 to 30th April 2022. In each panel, the dashed horizontal line shows the previous peak reached during the January 2021 wave in England; the shaded areas show 50% and 90% quantiles across each day, while the small triangles demarcate peaks from each of 50 individual model runs reached between 1st December 2021 and 30th April 2022. Numbers in overlays show the median and 95% projection intervals for total hospital admissions and deaths between 1st December 2021 and 30th April 2022. **(b)** Optimistic scenario (low immune escape, high booster efficacy) with Step 2 intervention. **(c)** Optimistic scenario with Step 1 intervention. **(d)** Optimistic scenario with Step 0 intervention. **(e)** Pessimistic scenario (high immune escape, low booster efficacy) with Step 2 intervention. **(f)** Pessimistic scenario with Step 1 intervention. **(g)** Pessimistic scenario with Step 0 intervention. **(h)** Same as panel f, but with Step 1 introduced seven days earlier on 26th December 2021. **(i)** Same as panel f, but with a greater reduction in transmission (15% overall) associated with “Plan B” measures introduced on 12th December 2021 than in panel f.

Under the optimistic scenario, our model projects that Step 2 measures reduce hospitalisations by 53,000 and deaths by 7,600 (**Fig. 3b**); that Step 1 measures reduce hospitalisations by 91,400 and deaths by 13,400 (**Fig. 3c**); and that Step 0 measures reduce hospitalisations by 104,900 and deaths by 15,240 (**Fig. 3d**) between 1st December 2021 and 30th April 2022 in England, compared with the current “Plan B” policy. Under the pessimistic scenario, Step 2 reduces hospitalisations by 160,000 and deaths by 23,800 (**Fig. 3e**); Step 1 reduces hospitalisations by 282,000 and deaths by 41,800 (**Fig. 3f**); and Step 0 reduces hospitalisations by 320,000 and deaths by 47,800 (**Fig. 3g**) between 1st December 2021 and 30th April 2022 in England, compared with the current “Plan B” policy. In general, the models project a larger exit wave for those scenarios with more stringent control measures enacted during December 2021–April 2022.

Under the pessimistic scenario, if Step 1 is introduced on 26th December 2021 instead of on 2nd January 2022 (7 days earlier), the model projects a median decrease of 21,000 hospitalisations and 4,200 deaths (**Fig. 3h**) between 1st December 2021 and 30th April 2022. If the “Plan B” measures which are introduced on 12th December 2021 have a 50% stronger impact than we previously estimated (22), the model projects a median decrease of 28,000 hospitalisations and 5,000 deaths (**Fig. 3i**) between 1st December 2021 and 30th April 2022.

We explored a number of scenarios varying the speed of the booster vaccination rollout, uptake of booster vaccinations, and the number of Omicron infections introduced to each NHS England region per day. Reducing the rate at which Omicron infections are introduced into England delays the projected epidemic and reduces the overall burden (**Fig. 4a–c**). Increases in the uptake of booster vaccinations substantially decrease the projected number of hospitalisations and deaths, emphasising the importance of the booster vaccination programme (**Fig. 4d–f**). Finally, while our base case scenario assumes a booster rollout of 500,000 vaccinations per day in England from 15th December 2021, we considered alternative assumptions of 200,000 and 350,000 booster vaccinations per day from the same date; slower booster vaccination rollout speeds do not substantially increase hospitalisations and deaths (**Fig. 4a**). This is because our model assumes that boosters are preferentially given to older individuals first, which means that by the time the Omicron variant is spreading rapidly, the most vulnerable individuals have already received their booster vaccines.

**Figure 4.**
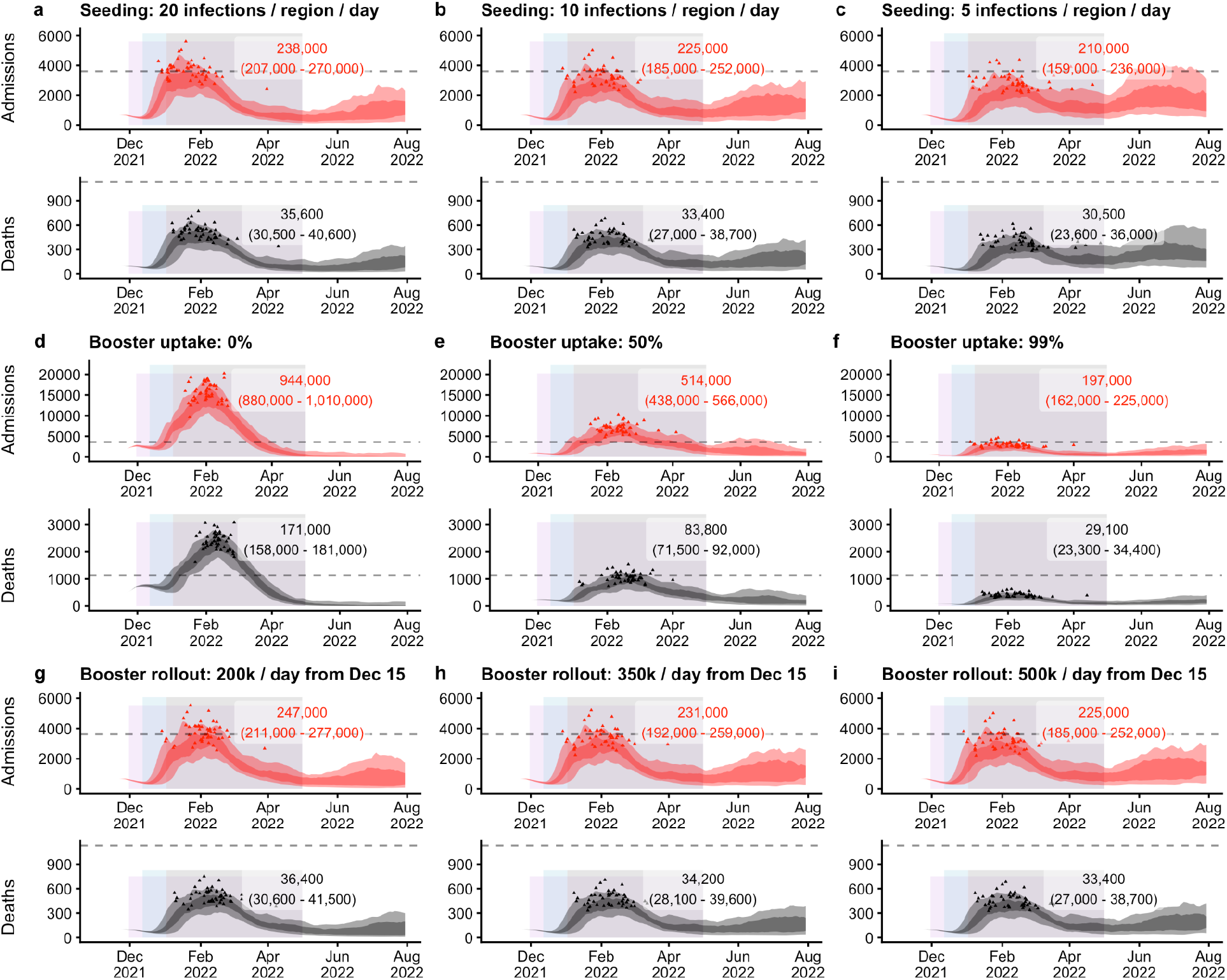
Sensitivity analyses. Daily hospital admissions and deaths projected for an Omicron epidemic in England, with increased mask-wearing from 30th November 2021 (lavender shaded rectangles), “Plan B” measures from 12th December 2021 (blue shaded rectangles), and Step 2 from 2nd January 2022 (grey shaded rectangles), with control measures in place until 30th April 2022. All sensitivity analyses assume the low immune escape, low booster efficacy scenario. In each panel, the dashed horizontal line shows the previous peak reached during the January 2021 wave in England; the shaded areas show 50% and 90% quantiles across each day, while the small triangles demarcate peaks from each of 50 individual model runs reached between 1st December 2021 and 30th April 2022. Numbers in overlays show the median and 95% projection intervals for total hospital admissions and deaths between 1st December 2021 and 30th April 2022. **(a–c)** Impact of initial seeding rate: **(a)** 20 infections, **(b)** 10 infections, or **(c)** 5 infections introduced into each NHS England region per day from 16th November 2021, for 20 consecutive days. **(d–f)** Impact of booster uptake of **(d)** 0%, **(e)** 50%, or **(f)** 99% in individuals 18 years or older who have completed their two-dose primary course. **(g–i)** Impact of booster rollout of **(g)** 200,000 booster vaccinations per day, **(h)** 350,000 booster vaccinations per day, or **(i)** 500,000 booster vaccinations per day in England from 15th December 2021, with average of 229,000 booster vaccinations per day beforehand. 500,000 boosters per day is our baseline assumption for other scenarios.

## Conclusions and discussion

These results suggest that the introduction of the Omicron B.1.1.529 variant in England will lead to a substantial increase in SARS-CoV-2 transmission, which, in the absence of strict control measures, has the potential for substantially higher case rates than those recorded during the Alpha B.1.1.7 winter wave in 2020–2021. This is due to Omicron’s apparent high transmissibility and ability to infect individuals with existing immunity to SARS-CoV-2 from prior infection or from vaccination (9) Our assumptions regarding the extent to which Omicron might evade the immune response are in line with existing knowledge of previous VOCs’ neutralisation, early neutralisation studies of Omicron and preliminary vaccine efficacy estimates (1–6,21). The majority of scenarios considered project that without the implementation of further control measures, hospital admissions resulting from the Omicron wave of transmission could exceed the peak levels recorded in England during the previous winter wave in 2020-2021. Additional control measures may therefore be required to minimise disease burdens and to protect healthcare services.

We estimate the growth rate of symptomatic SGTF cases in England as a proxy for the growth rate of Omicron itself. These estimates suggest a 2.4 day doubling time. For the higher immune escape scenario (a 12.8-fold drop in neutralisation titre relative to the Delta B.1.617.2 variant), we estimate that Omicron is 5–10% less transmissible than Delta. For the lower immune escape scenario (a 5.1-fold drop in neutralisation titre relative to the Delta B.1.617.2 variant), we estimate that Omicron is 30–35% more transmissible than Delta.

For our most optimistic scenario (low immune escape and highly effective booster vaccines), an Omicron epidemic without the introduction of additional control measures may not exceed the peak levels of hospitalisations recorded in January 2021. However, for our most pessimistic scenario (high immune escape and less effective booster vaccines), we project that hospitalisations and potentially deaths will exceed the peak levels recorded in January 2021.

The introduction of control measures is projected to partially suppress Omicron transmission; however, in the most pessimistic scenario we project that stringent control measures such as those implemented following the Alpha B.1.1.7 winter wave of transmission may be required to ensure that healthcare services are not overwhelmed. These results suggest that increased uptake of COVID-19 booster vaccinations can mitigate the projected burden. In our model, increasing the speed at which booster vaccines are administered has little effect on projected outcomes, since our model assumes that boosters are administered strictly in decreasing order of age and hence that the majority of vulnerable individuals in England have already received booster vaccinations by the time of the Omicron wave. We stress that in the real world, if an increased speed of booster rollout can help reach more vulnerable individuals before Omicron infections surge, there would likely be a much more substantial benefit of increasing the speed of booster rollout than we have been able to capture here. There remains significant uncertainty about the timing of projected epidemic peaks as well as the time at which control measures may need to be implemented. We have shown that small changes in the number of Omicron introductions per day early in the epidemic can shift the projected epidemic burden later, allowing more time for control measures to be taken. However, measures to reduce Omicron introductions become comparatively less important once the variant has spread substantially within the country.

Our work is subject to limitations. We do not account for the differing rate of Omicron introduction to each NHS England region, and we do not consider the impact of localised interventions. We do not capture the potential impact of newly available antiviral therapies, or the future availability of targeted vaccines for Omicron, but nor do we consider that some existing therapies (such as monoclonal antibodies) may become less effective given the escape properties of Omicron. We assume that the baseline infection fatality rate (IFR) remains constant over the projection period (except as altered by, e.g., primary and/or booster vaccination uptake), though our model fit suggests the IFR may increase during periods of high strain on hospital services (**Fig. S1**). We only report burdens during the period 1st December 2021 to 30 April 2022, because of uncertainty over what additional measures may be available to mitigate Omicron by mid-2022—for example, reformulated vaccines. However, our scenarios with more stringent control measures enacted between December 2021 to April 2022 result in larger exit waves after control measures are lifted. Finally, the control measures we consider are limited and are based on the known impacts of previously implemented control strategies for SARS-CoV-2 in England. Implementing strategies such as enhanced mass testing may help to reduce the required stringency of non-pharmaceutical interventions aimed at reducing interpersonal contact rates. Accordingly, there is an urgent need to rapidly assess the feasibility and potential impact of such alternative control strategies.

There is still substantial uncertainty surrounding the biological characteristics of the Omicron variant (particularly its clinical severity), the effectiveness of existing vaccines and pharmaceuticals, and the efficacy of control measures enacted by policymakers for suppressing SARS-CoV-2 transmission. In populations with high levels of immunity such as England and South Africa, it is clear that the Omicron variant has the potential to cause significant disruption, particularly if it exhibits higher levels of immune escape. It remains unclear whether Omicron will outcompete pre-existing variants in other settings with lower levels of existing immunity in the event that its inherent transmissibility is lower than that of the currently-dominant Delta variant. The consequences of Omicron will become clearer as more evidence emerges.

## Data Availability

Data sources used for these analyses are either publicly available, or were provided to RCB and NGD as members of the UK's Scientific Pandemic Influenza Modelling (SPI-M) group, which provides expert advice to the UK's Department of Health and Social Care and wider UK government on scientific matters. Specific permissions have been sought to use SPI-M data for this publication. The SPI-M datasets used for model fitting are unpublished and not publicly available, but are closely aligned with the UK Government's COVID-19 dashboard (see https://coronavirus.data.gov.uk/) and other publicly available sources such as the Wellcome Sanger Institute's COVID-19 genomic surveillance data (see https://covid19.sanger.ac.uk/downloads).

## Author contributions

RCB and NGD accessed and verified the data and conducted the analyses. All authors contributed to study design and drafting of the manuscript.

## Statement on data availability

Data sources used for these analyses are either publicly available, or were provided to RCB and NGD as members of the UK’s Scientific Pandemic Influenza Modelling (SPI-M) group, which provides expert advice to the UK’s Department of Health and Social Care and wider UK government on scientific matters. Specific permissions have been sought to use SPI-M data for this publication. The SPI-M datasets used for model fitting are unpublished and not publicly available, but are closely aligned with the UK Government’s COVID-19 dashboard (12) and other publicly available sources such as the Wellcome Sanger Institute’s COVID-19 genomic surveillance data (28).

## Statement on code availability

Analysis and code will be made available upon publication.

## Ethics

Ethical approval for this research was given by the London School of Hygiene & Tropical Medicine Ethics Committee, project ID: 22828.

## Supplementary material

**Figure S1.**
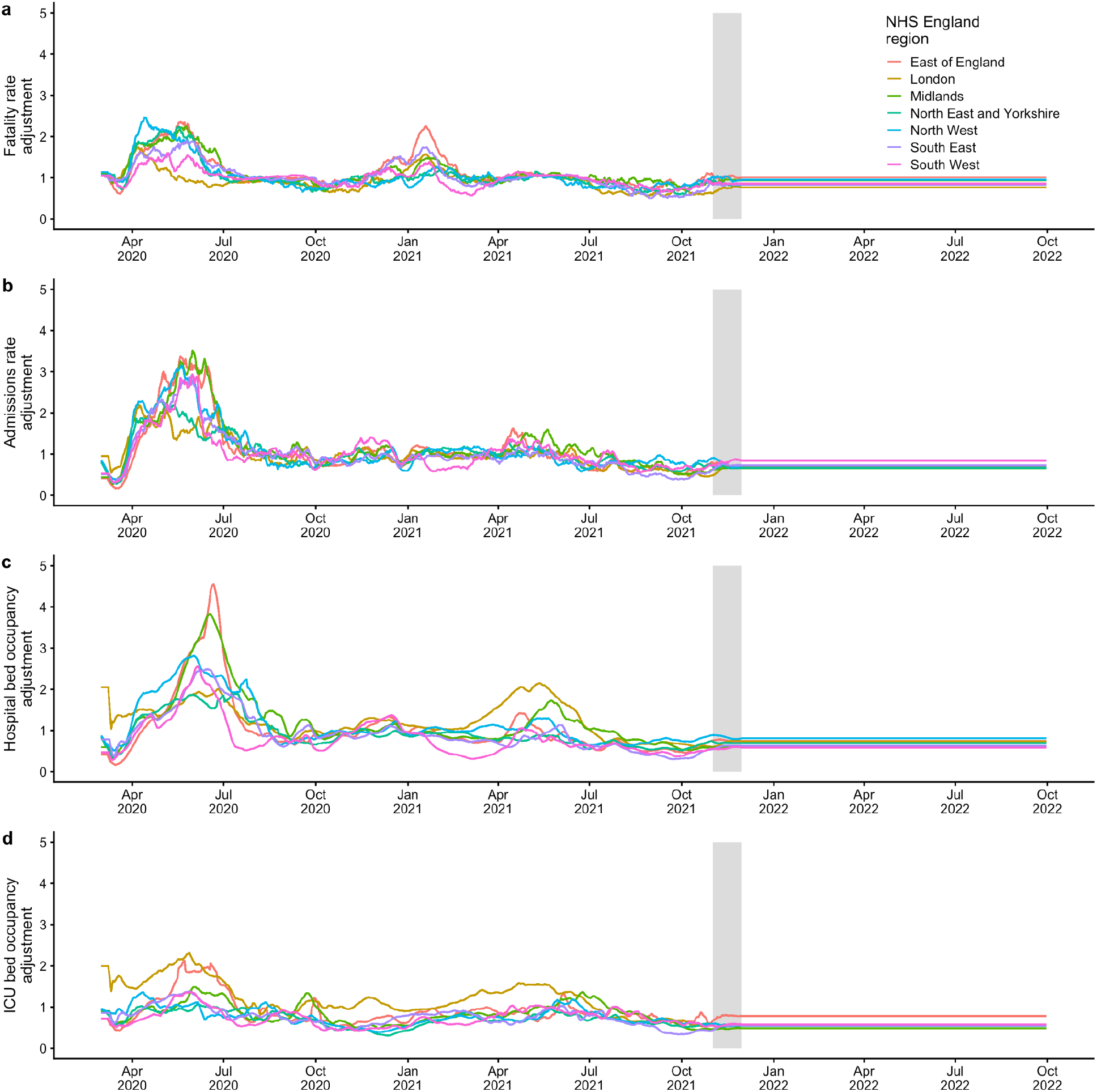
**(a)** Modelled adjustments to the infection fatality rate over time, plotted for all seven NHS England regions, for the low booster efficacy scenario. **(b)** Modelled adjustments to the hospital admission rate over time. **(c)** Modelled adjustments to the hospital bed occupancy rate over time. **(d)** Modelled adjustments to the intensive care unit (ICU) bed occupancy rate over time. NHS = National Health Service.

**Figure S2.**
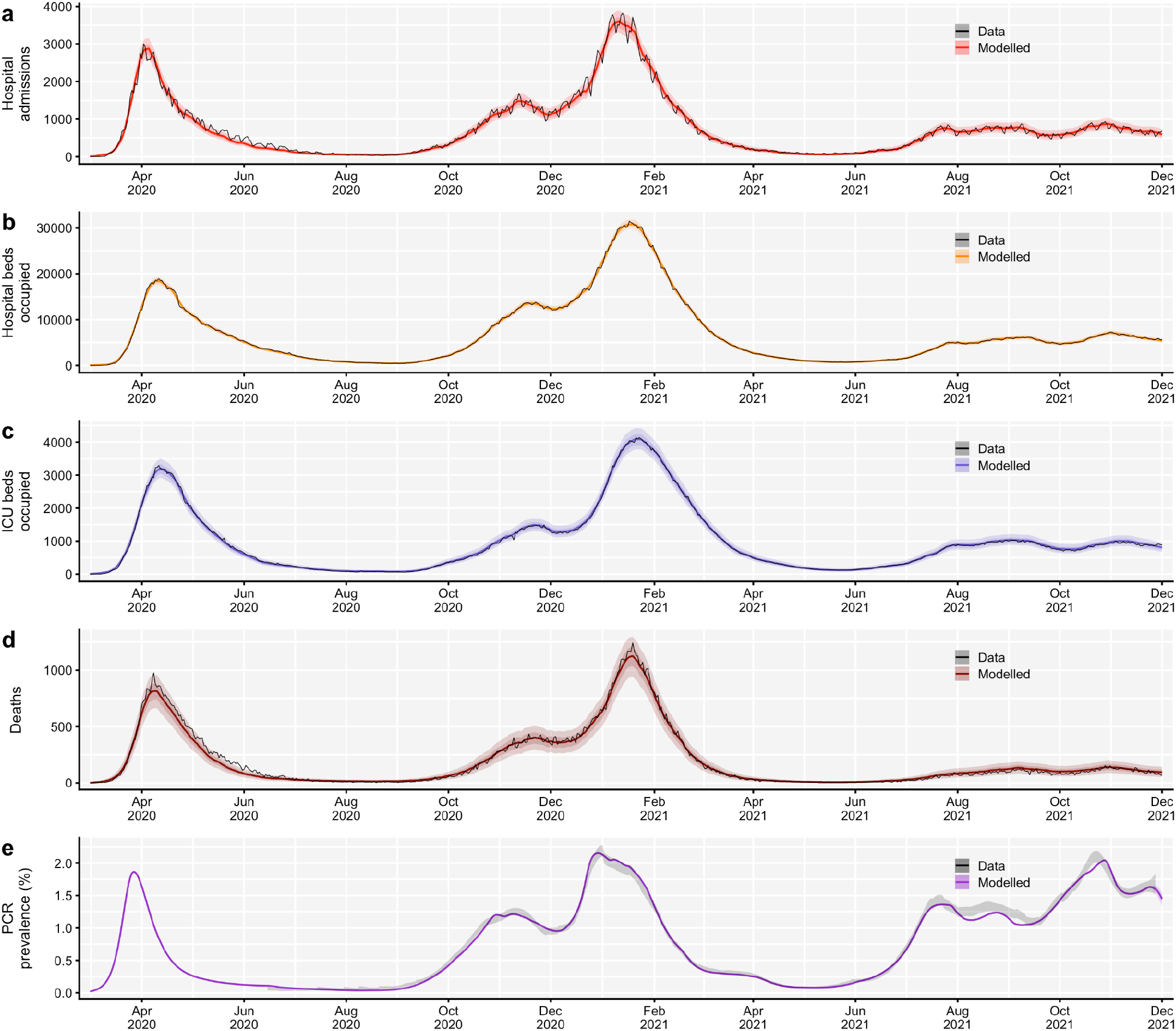
Comparison of fitted transmission model to data for England. Transmission model comparison to data in England, March 2020 – November 2021, for **(a)** hospital admissions, **(b)** hospital beds occupied, **(c)** ICU beds occupied, **(d)** deaths, and **(e)** PCR prevalence.

